# Cancer treatment patterns and factors affecting receipt of treatment in older adults: results from the ASPREE Cancer Treatment Substudy (ACTS)

**DOI:** 10.1101/2022.11.01.22281781

**Authors:** Jaidyn Muhandiramge, Erica T. Warner, John R. Zalcberg, Andrew Haydon, Galina Polekhina, G J. van Londen, Peter Gibbs, Wendy B. Bernstein, Jeanne Tie, Jeremy L. Millar, Victoria J. Mar, John J. McNeil, Robyn L. Woods, Suzanne G. Orchard, ASPREE Investigator Group

## Abstract

Cancer treatment planning in older adults is complex and requires careful balancing of survival, quality of life benefits, and risk of treatment-related morbidity and toxicity. As a result, treatment selection in this cohort tend to differ from younger patients. However, there are very few studies describing cancer treatment patterns in older cohorts. We used data from the ASPirin in Reducing Events in the Elderly (ASPREE) trial and the ASPREE Cancer Treatment Substudy (ACTS) to describe cancer treatment patterns in the elderly. We used a multivariate logistic regression model to identify factors affecting receipt of treatment. Of 1,893 eligible Australian and United States (US) participants with incident cancer, 1,569 (81%) received some form of cancer treatment. Non-metastatic breast cancers most frequently received treatment (98%), while haematological malignancy received the lowest rates of treatment (60%). Factors associated with not receiving treatment were older age (OR 0.94, 95% CI 0.91-0.96), residence in the US (OR 0.35, 95% CI 0.22-0.56), smoking (OR 0.60, 95% CI 0.37-0.98), and diabetes (OR 0.58, 95% CI 0.41-0.82). After adjustment for treatment patterns in sex-specific cancers, sex did not impact receipt of treatment. This study is one of the first describing cancer treatment patterns and factors affecting receipt of treatment across common cancer types in older adults. We found that most older adults with cancer received some form of cancer treatment, typically surgery or systemic therapy, although this varied with factors including cancer type, age, sex, and country of residence.

## Introduction

Cancer is one of the most common diseases worldwide, and a leading cause of morbidity and death.^1^ The incidence of cancer in older adults is increasing, driven predominantly by increasing numbers of individuals living into old age.^1^ An increase in the availability of screening and surveillance, coupled with the development of newer, more effective therapies has led to a decrease in mortality across several cancer types.^1^ In Australia, cancer survivor numbers are estimated to grow from 1.1 million in 2018 to 1.9 million in 2040,^2^ and in the United States (US), an additional 5.2 million survivors are anticipated between 2019 and 2030.^3^

Cancer management in older adults is complex, with many factors determining whether treatment is offered including patient age, frailty, and comorbidities, along with the goals of patient care.^4, 5^ In a cohort where life expectancy is limited irrespective of cancer, older adults with cancer are typically less likely to be offered aggressive treatment.^6^ In those who do receive treatment, careful selection of treatment modality and regimen is required to avoid treatment-related toxicity and morbidity.^6^

The elderly represent almost half of the global cancer population, but this group is frequently underrepresented in cancer treatment clinical trials.^7^ Similarly, while several studies have investigated cancer treatment patterns, these are usually limited to cohorts of younger patients, often those in late middle-age.^8, 9^ Furthermore, these studies typically focus on a single, or few, cancer types and often do not analyse factors affecting receipt of treatment.

We used data from the ASPirin in Reducing Events in the Elderly (ASPREE) trial which captured cancer events and associated data, along with treatment data collected within the ASPREE Cancer Treatment Substudy (ACTS), to explore the characteristics of older persons receiving cancer treatment and the types of treatment employed. We aim to describe cancer treatment patterns in older adults diagnosed with cancer and explore factors affecting receipt of cancer treatment.

## Methods

### Study design

ACTS was designed as a substudy nested within the ASPREE trial. ASPREE was a randomised, double-blinded, placebo-controlled, multi-institution clinical trial investigating whether daily low dose (100mg) aspirin prolonged disability free survival in healthy older people. The study protocol and baseline participant characteristics of ASPREE have been previously published,^10, 11^ along with the main trial findings.^12-14^

### Study population

The ASPREE trial recruited eligible participants from 2010-2014 and collected data via annual in-person visits and regular phone calls, for a median 4.7 years. A total of 19,114 community-dwelling individuals aged ≥70 years (≥65 if a US minority) from both Australia and the US were enrolled. To be eligible, participants needed to be free from cardiovascular disease, dementia, and significant physical disability. Prior history of cancer was not an exclusion criteria (19% had a past cancer history),^15^ but eligible participants had to be free from a disease likely to cause death within 5 years.

ASPREE participants from both countries, with an incident cancer event during the trial, were included in ACTS. Participants with only a history of cancer pre-randomisation (i.e. no in-trial cancer) were not included. However, past cancer did not preclude participants from inclusion in ACTS if an in-trial event occurred. ACTS attempted to collect cancer treatment data for the 1,933 participants diagnosed with incident cancer during ASPREE.

### ASPREE cancer adjudication

The details of cancer event capture and adjudication for ASPREE are described elsewhere.^16^ Briefly, ASPREE captured in-trial cancer events through participant self–report and review of medical records. These data included cancer type, date of diagnosis or date metastatic disease was discovered, stage, and type of event (*i.e*. metastatic *versus* non-metastatic). Clinical documentation and investigations relating to the event were assigned to two clinical experts to confirm or refute the report based on pre-specified criteria. If the two adjudicators were not in accordance, a third adjudicator would review the case. For participants with pre-randomisation cancers, any post-randomisation cancer needed to be a new cancer or development of metastatic disease of a pre-existing cancer.

### ACTS data collection

Cancer treatment data were extracted from the supporting documents sourced during ASPREE for event adjudication, or from a participant’s specialist. Data were collected to define whether a participant had received cancer treatment, the cancer for which it was prescribed, and the modality of treatment. A participant’s treatment status was only listed as “No treatment” if there was definitive evidence that they had not received treatment. If treatment status for any modality was unclear, they were excluded from the analysis. Treatment categories and coding rules were developed in consultation with at least two practicing medical oncologists and are described in Supplementary Table 1. Data on treatment dose, duration, line, intent, or regimen were not collected. Treatment data were only collected for cancer events that occurred during the ASPREE time period (*i.e*. prior to 12 June 2017). For participants who had two cancer events for the same cancer types (*e.g*. a non-metastatic breast and a metastatic breast endpoint), the same treatment regime was entered for both events. If a participant developed a different type of cancer, then the treatment for each cancer type was evaluated independently.

### Statistical analysis

Statistical analysis was conducted in R (R Core Team, 2014). Participant characteristics between “Treatment” and “No treatment” groups were compared using a two-sample t-test or chi-square test for continuous and categorical variables, respectively. Multivariate logistic regression was used to assess the association between baseline factors and receipt of cancer treatment. Only variables deemed to be clinically relevant were included in the models. The linearity assumption was assessed for continuous variables by inspecting a plot between each predictor and the logit values. Collinearity was assessed by calculating variance inflation factors and was not present between variables.

## Results

Baseline characteristics of the ACTS cohort are summarised in Table 1. Of the 1,933 ASPREE participants with post-randomisation cancer, 1,893 ASPREE participants were included in ACTS (median age 74.58; 56% male; 94% Non-Hispanic Caucasian; 91% Australian). Median age, race, and country of residence of ACTS participants were similar to that of the total ASPREE cohort, although a greater proportion of ACTS participants were male compared to ASPREE (44% male). Of the ACTS cohort, 1,569 (83%) had received some form of cancer treatment. Nearly four-fifths consumed alcohol, while only a very small minority were current smokers. A slightly greater proportion of ACTS participants were current or former smokers compared to ASPREE participants. Similar to the ASPREE cohort, hypertension and dyslipidaemia were prevalent in the ACTS cohort (76% and 63% respectively), while a smaller proportion had chronic kidney disease or diabetes (32% and 13% respectively). Diabetes was more prevalent in those who did not receive cancer treatment. Almost all participants (98%) were not frail upon entry to ASPREE. A flow diagram illustrating ACTS eligibility can be found in Supplementary Figure 1.

**Table 1.** Participant demographics and characteristics of the total cohort, as well as stratification by incident (first) post-randomisation cancer diagnosis and receipt of cancer treatment.

|  | Total ASPREE cohort<br>n = 19,114<br>(% of column) | ACTS cohort |  |  | P Value<br>(No treatment<br>versus Treatment) |
| --- | --- | --- | --- | --- | --- |
|  |  | Total<br>n = 1,893<br>(% of column) | No cancer treatment<br>n = 324<br>(% of column; % of<br>row) | Cancer treatment<br>n = 1,569<br>(% of row; % of<br>column) |  |
| Age at randomisation [years;<br>median (Q1-Q3)] | 74.0<br>(71.6-77.7) | 74.6 (72.0-78.7) | 76.2<br>(72.4-80.5) | 74.45<br>(71.9-78.2) | < 0.001 |
| Age at cancer diagnosis [years;<br>median (Q1-Q3)] | N/A | 77.3<br>(74.6-81.4) | 78.87<br>(75.1-84.0) | 77.14<br>(74.5-80.9) | < 0.001 |
| Sex |  |  |  |  |  |
| Male | 8,332 (44%) | 1,053 (56%) | 210 (65%, 20%) | 843 (54%, 80%) | < 0.001 |
| Female | 10,782 (56%) | 840 (44%) | 114 (35%, 14%) | 726 (46%, 86%) |  |
| Race |  |  |  |  |  |
| Non-Hispanic Caucasian | 17,449 (91%) | 1,770 (94%) | 303 (93%, 17%) | 1,469 (94%, 83%) | 0.359 |
| Non-Hispanic Asian | 164 (1%) | 13 (1%) | 1 (<1%, 8%) | 12 (1%, 92%) |  |
| Non-Hispanic African<br>American | 901 (5%) | 56 (3%) | 14 (4%, 25%) | 42 (3%, 75%) |  |
| Non-Hispanic Other | 111 (<1%) | 18 (1%) | 4 (1%, 22%) | 14 (1%, 78%) |  |
| Hispanic | 488 (3%) | 35 (2%) | 4 (1%, 11%) | 31 (2%, 89%) |  |
| Country |  |  |  |  |  |
| Australia | 16,703 (87%) | 1,718 (91%) | 274 (85%, 16%) | 1,444 (92%, 84%) | < 0.001 |
| United States | 2,411 (13%) | 175 (9%) | 50 (15%, 29%) | 125 (8%, 71%) |  |
| Rurality<sup>a</sup> |  |  |  |  |  |
| Major cities | 8,729 (52%) | 947 (50%) | 136 (50%, 14%) | 811 (56%, 86%) | 0.084 |
| Inner regional | 5,976 (36%) | 587 (31%) | 100 (37%, 17%) | 487 (34%, 83%) |  |
| Outer regional | 1,947 (12%) | 180 (9%) | 37 (14%, 21%) | 143 (10%, 79%) |  |
| IRSAD percentile<sup>b</sup> | 58 (31-83) | 59 (34-85) | 60 (35-85) | 57 (30-82) | 0.088 |
| Education |  |  |  |  |  |
| ≤12 years of education | 10,955 (57%) | 1,088 (57%) | 194 (60%, 18%) | 894 (57%, 82%) | 0.403 |
| 13-15 years of education | 3,255 (17%) | 345 (18%) | 50 (15%, 14%) | 295 (19%, 86%) |  |
| ≥16 years of education | 4,903 (26%) | 460 (24%) | 80 (25%, 17%) | 380 (24%, 83%) |  |
| BMI<sup>c</sup> |  |  |  |  |  |
| Underweight (<23) | 2,194 (12%) | 182 (10%) | 32 (10%, 18%) | 150 (10%, 82%) | 0.771 |
| Normal weight (24-30) | 11,224 (59%) | 1,150 (61%) | 201 (62%, 17%) | 949 (61%, 83%) |  |
| Overweight (≥30) | 5,607 (30%) | 553 (29%) | 90 (28%, 16%) | 463 (30%, 84%) |  |
| Alcohol use |  |  |  |  |  |
| Current | 14,642 (77%) | 1,492 (79%) | 251 (78%, 17%) | 1,241 (79%, 83%) | 0.681 |
| Former | 1,136 (6%) | 125 (6%) | 25 (8%, 20%) | 100 (6%, 80%) |  |
| Never | 3,336 (18%) | 276 (15%) | 48 (15%, 17%) | 228 (15%, 83%) |  |
| Smoking status |  |  |  |  |  |
| Current | 735 (4%) | 120 (6%) | 29 (9%, 23%) | 91 (6%, 77%) | 0.081 |
| Former | 7,799 (41%) | 850 (45%) | 147 (45%, 17%) | 703 (45%, 83%) |  |
| Never | 10,580 (55%) | 923 (49%) | 148 (46%, 16%) | 775 (49%, 84%) |  |
| Chronic disease |  |  |  |  |  |
| Chronic kidney disease | 4,740 (25%) | 573 (30%) | 98 (32%, 17%) | 475 (32%, 83%) | 1.000 |
| Diabetes | 2,045 (11%) | 242 (13%) | 65 (20%, 27%) | 177 (11%, 73%) | < 0.001 |
| Dyslipidaemia | 12,467 (65%) | 1,195 (63%) | 200 (61%, 17%) | 995 (63%, 83%) | 0.522 |
| Hypertension | 14,195 (74%) | 1,437 (76%) | 252 (77%, 18%) | 1,185 (76%, 82%) | 0.542 |
| Frailty<sup>d</sup> |  |  |  |  |  |
| Not frail | 11,246 (59%) | 1,066 (56%) | 163 (50%, 15%) | 903 (58%, 85%) | 0.033 |
| Pre-frailty | 7,447 (39%) | 779 (41%) | 149 (46%, 19%) | 630 (40%, 81%) |  |
| Frailty | 421 (2%) | 48 (3%) | 12 (4%, 25%) | 36 (2%, 75%) |  |
| Polypharmacy | 5,088 (27%) | 506 (27%) | 101 (31%, 20%) | 405 (26%, 80%) | 0.064 |
| Aspirin intervention | 9,589 (50%) | 930 (49%) | 163 (50%, 18%) | 767 (49%, 82%) | 0.760 |

Abbreviations: BMI = Body Mass Index, IRSAD = Index of Relative Socio-economic Advantage and Disadvantage.
NB: Some percentages may not add to 100% where there are participants with missing data.
<sup>a</sup> Australian participants only.
<sup>b</sup> A summary measure of economic and social conditions within an area. A low score corresponds to greater disadvantage. Here, we present percentiles for ease of interpretation.
<sup>c</sup> BMI as per recommendations for the elderly (>65 years).
<sup>d</sup> As classified using the Fried Frailty Index.

The characteristics of the cancer treatment received by the ACTS cohort are outlined in Table 2. The most common type of cancer treatment was surgery (54% of those receiving treatment), followed by systemic therapy (46%), radiation therapy (29%), and regional therapy^a^ (1%). Usually only one major treatment modality was administered (55%), but among those receiving multimodal therapy, systemic therapy plus surgery (28% of those receiving treatment) was the most common. The most common form of systemic therapy was cytotoxic chemotherapy (62% of all systemic therapies), with usually only one therapy administered (85%).

**Table 2.** Characteristics of the incident cancer diagnosed following randomisation and their treatment in participants who received a post-randomisation cancer diagnosis during ASPREE.

|  | Cancer treatment cohort (n = 1,893) |
| --- | --- |
| **No cancer treatment<sup>a</sup>** | 324 (17%) |
| **Any cancer treatment<sup>a</sup>** | 1,569 (83%) |
| Systemic therapy | 869 (46%) |
| Cytotoxic chemotherapy | 537 (28%) |
| Hormonal therapy | 351 (19%) |
| Targeted therapy | 85 (5%) |
| Immunotherapy | 31 (2%) |
| Radiation therapy | 544 (29%) |
| Surgery | 1,029 (54%) |
| Regional therapy | 16 (1%) |
| **Combination therapy<sup>b</sup>** |  |
| Systemic therapy and surgery | 435 (28%) |
| Systemic therapy and radiation therapy | 368 (23%) |
| Radiation therapy and surgery | 266 (17%) |
| Radiation therapy, systemic therapy, and surgery | 188 (12%) |
| **Number of major treatment modalities<sup>b,c</sup>** |  |
| Only one modality | 868 (55%) |
| Two modalities | 505 (36%) |
| Three modalities | 188 (12%) |
| **Number of types of systemic therapy<sup>d</sup>** |  |
| Only one type | 741 (85%) |
| Two types | 121 (14%) |
| Three or more types | 7 (1%) |
| Percentages in some subgroups may not total to 100% as some participants received multiple modalities of cancer treatment. |  |
| <sup>a</sup> Percentage of total ACTS cohort (n = 1,893). |  |
| <sup>b</sup> Percentage of participants who received any cancer treatment (n = 1,569). |  |
| <sup>c</sup> The 8 missing participants in this group received “Regional therapy” and were not counted as having received a “Major treatment modality”. |  |
| <sup>d</sup> Percentage of participants who received systemic therapies (n = 869). |  |

Table 3 summarises the modalities of cancer treatment received by the six most common cancer types, along with the frequency of each cancer type in the ACTS cohort, and stratification of treatment by age, survival time, and cause of death. The most common cancer types diagnosed during ASPREE were blood, breast, colorectal, lung, prostate cancers, and melanoma. The majority of these were non-metastatic (n=937). Nearly all participants with non-metastatic breast cancer received treatment (98%), while those with non-metastatic prostate and haematological cancers were least likely to receive treatment (69% and 60% of the time, respectively). Those with metastatic breast, colorectal and prostate cancer received treatment in similarly high proportions (88%). Generally, older participants received treatment less frequently than younger participants; those aged 70-75 years (85%) received the greatest amount of treatment, and this held across all treatment modalities. Participants with greater time from cancer diagnosis to death (i.e. survival time) more frequently received cancer treatment. Treatment data for the less common cancer types can be found in Supplementary Table 2. Treatment data stratified by sex and country can be found in Supplementary Tables 3 and 4, respectively.

**Table 3.**
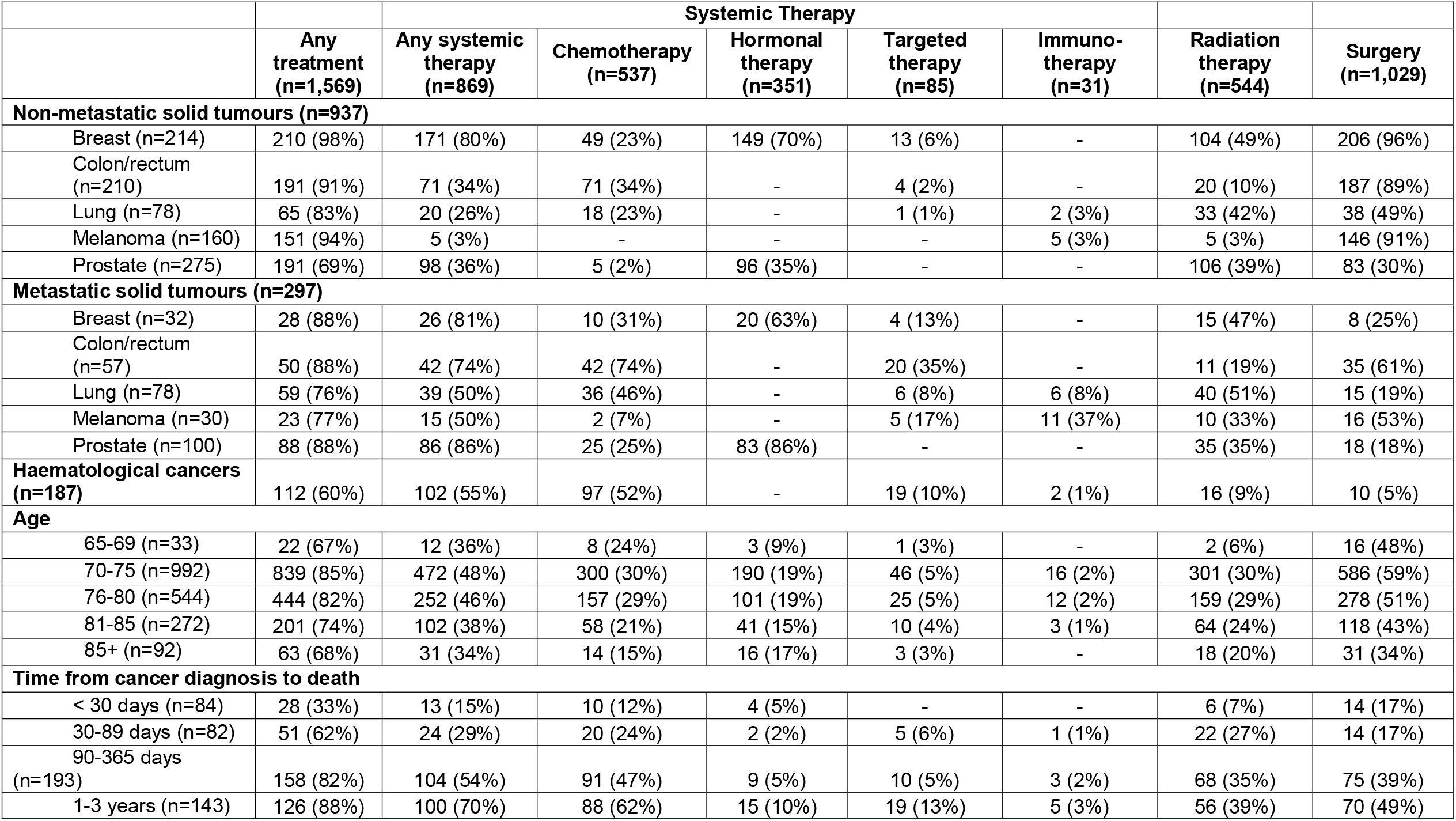
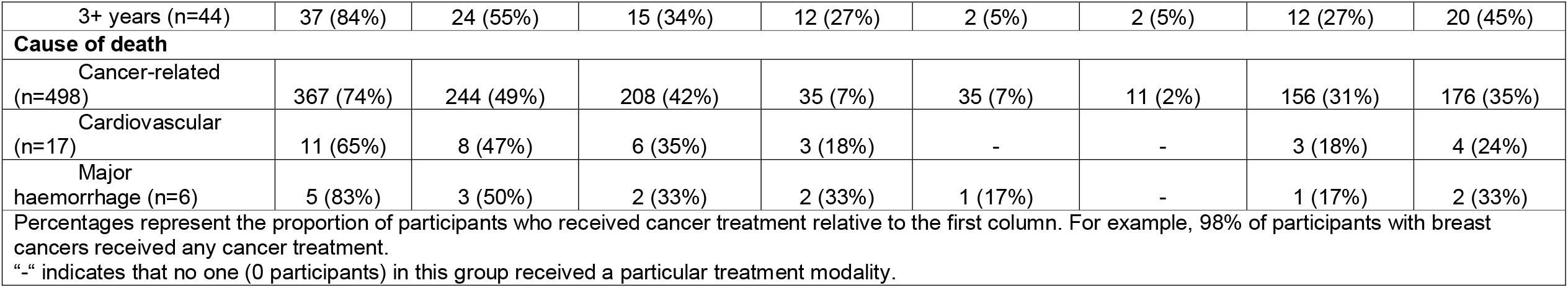
Cancer treatment modalities received for incident post-randomisation cancers during ASPREE, stratified by most common cancer types and metastatic status, and cause of death and time from diagnosis to death.

The impact of various baseline factors on the receipt of cancer treatment was assessed using a multivariate logistic regression model. In summary, increasing age (OR 0.94, 95% CI 0.91-0.96), residence in the US (OR 0.35, 95% CI 0.22-0.56), current smoking status (OR 0.60, 95% CI 0.37-0.98), and diabetes (OR 0.58, 95% CI 0.41-0.82) reduced the likelihood of receiving any treatment. Sensitivity analysis excluding non-Caucasian participants (OR 0.37 95% CI 0.23-0.61) and African American participants (OR 0.40, 95% CI 0.26-0.62) attenuated the impact of US residence but did not remove it entirely. Female sex was associated with receipt of treatment (OR 1.76, 95% CI 1.35-2.31), although on sensitivity analysis excluding sex-specific cancers, this association was no longer present (OR 1.05, 95% CI 0.77-1.44). The results of the models, including those analysing receipt of systemic therapy, radiation therapy, and surgery, are detailed in Figures 1 and 2.

**Figure 1.**
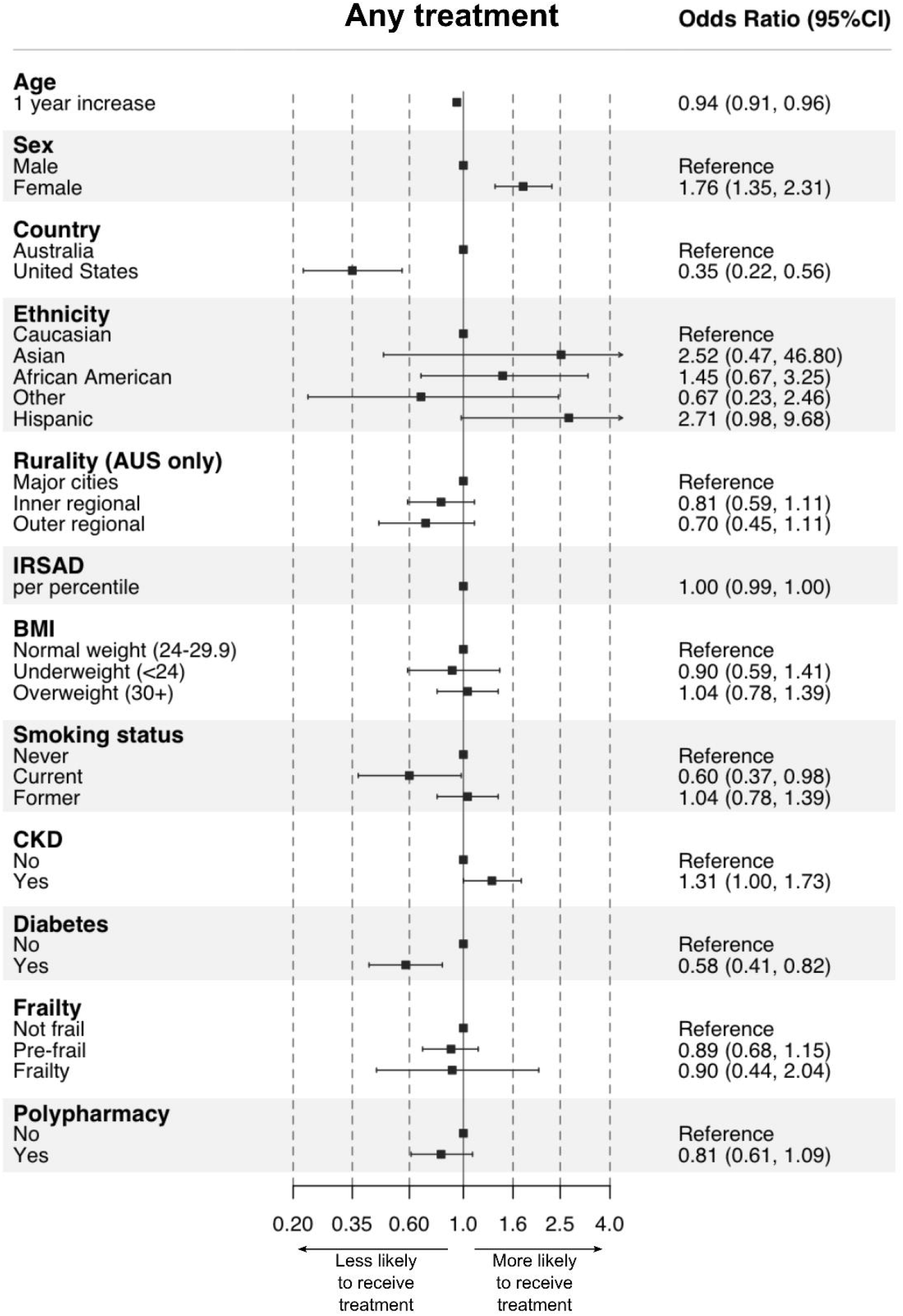
Associations between demographic factors, health behaviours, and chronic conditions and receipt of any cancer treatment for incident post-randomisation cancers during ASPREE.

**Figure 2.**
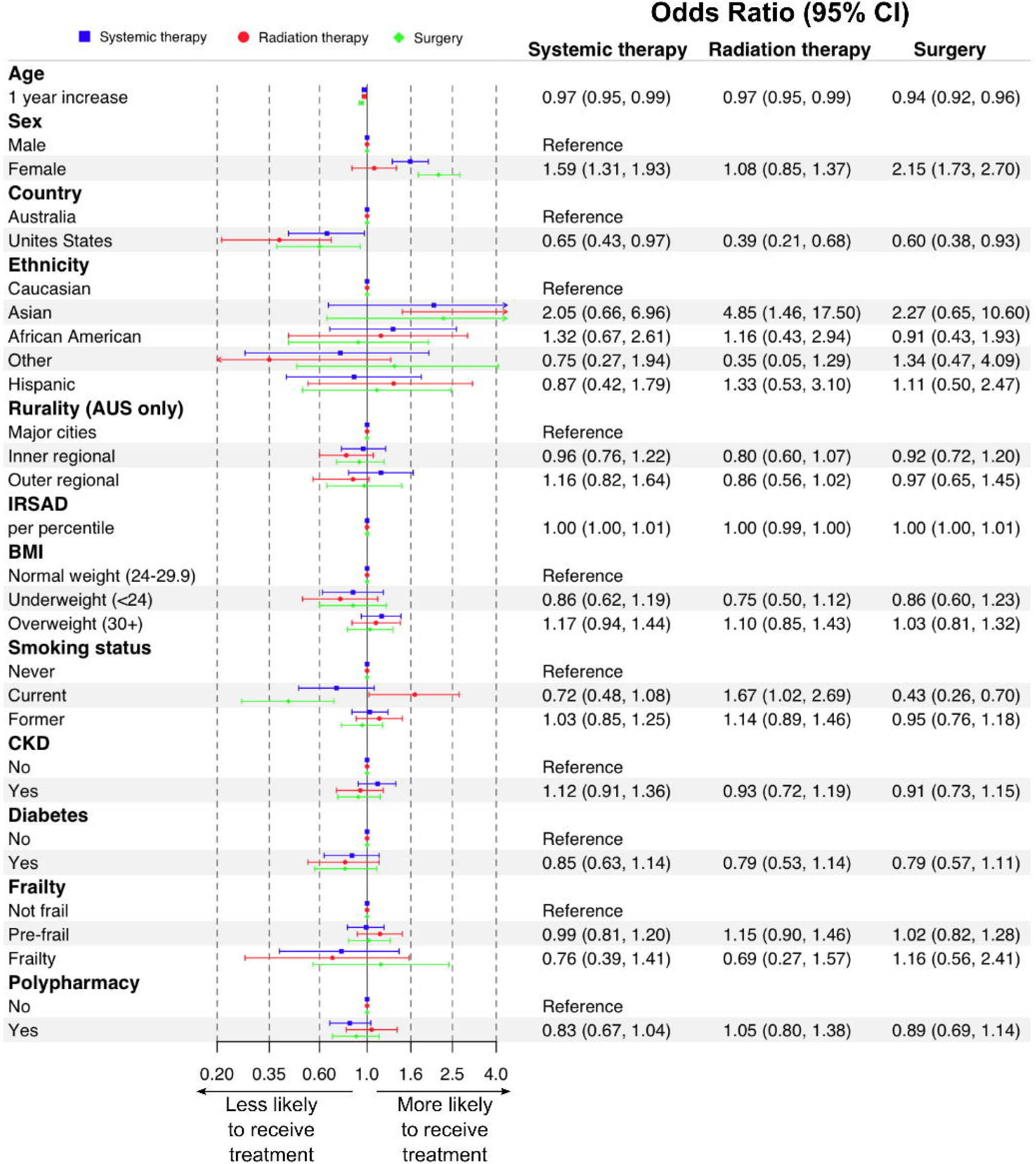
Associations between demographic factors, health behaviours, and chronic conditions and receipt of systemic therapy, radiation therapy, and surgery for incident post-randomisation cancers during ASPREE.^b^

## Discussion

The ASPREE Cancer Treatment Substudy (ACTS) adds valuable information to the literature regarding cancer treatment patterns in older adults with various types of cancer. In our cohort, most older but otherwise reasonably healthy adults with cancer received some form of cancer treatment, with roughly half receiving at least one of systemic therapy or surgery and younger participants more frequently receiving treatment. Slightly more female participants received treatment than males (86% versus 80%), while more Australian participants received treatment than US participants (84% versus 71%). Treatment rates generally increased with increasing survival time, although this is not surprising given that those with advanced disease and short life expectancies are unlikely to be offered treatment, particularly in cases where systemic therapy or surgery requires a reasonable functional baseline.^17^

Our treatment pattern data generally aligns with cancer data collected by the National Cancer Registration and Analysis Service in England, where 45% received surgery and 27% received radiation therapy. While the service’s 2020 report did not look at systemic therapy overall, it states that 28% received cytotoxic chemotherapy, in line with our cohort.^18^ Notably, the English data represents all patients with cancer, not just older adults. While some Australian cancer data exist in the form of National Cancer Control Indicators published by Cancer Australia,^19^ cancer treatment patterns are not widely available. Similarly, data from the US on overall cancer treatment patterns is not easily accessible, although Miller et al. presents a detailed analysis of site-specific treatment patterns using Surveillance, Epidemiology, and End Results (SEER) data.^20^ While overall treatment patterns provide a broad overview of how cancer in older adults is managed, disease-specific stratification is essential given the heterogeneity in treatments offered for various cancer types. In general, treatment rates for the six most common cancers in our study did not differ greatly from existing literature.

Participants with haematological malignancies received treatment in slightly greater rates than elderly cohorts with acute leukaemia in the US,^21, 22^ but in lower rates than US patients with slower growing haematological malignancies.^23^ As ASPREE grouped all types of haematological malignancy together, the heterogeneity of cancer aggressiveness, treatments offered, and stage may provide an explanation for this variation. Participants with haematological malignancy also received some of the lowest rates of treatment overall (60%), possibly due to a predominance of classically non-aggressive subtypes (*e.g*. chronic lymphocytic leukaemia) where a ‘watchful waiting’ approach is often employed.^20, 24^

For breast cancer, our data resemble published data from both Australia and the US where most patients with non-metastatic breast cancer undergo surgery.^20, 25, 26^ Rates of surgical treatment were greatest in this group, likely due to the use of relatively low-risk breast-conserving procedures. Our cohort received slightly less radiation therapy than that reported in a younger Australian cohort (median age 61 versus 74.6 years) where 63% received radiation therapy.^25^ Rates of radiation therapy were still the highest for the six commonest cancers, an expected finding given the widespread use of adjuvant radiation in localised or regional breast cancers.^20^ In those with metastatic disease, rates of chemotherapy (31%) and hormonal therapy (63%) resemble the rates of large European and North American cohorts.^9^ Here, hormone receptor status data can add nuance to treatment patterns, although that data were not available for our analysis.

We demonstrated similar treatment patterns for prostate cancer compared to published literature, where roughly one-third undergo surgery and a slightly higher proportion receive radiation,^27^ although younger Australian men are more likely to undergo radical prostatectomy (47% described by Wang et al.).^28^ Radiation was the most common treatment (39%) for early-stage prostate cancer, an expected finding given the similar efficacy^29^ and relative safety of radiation therapy versus surgery, particularly in our older cohort. ACTS participants with metastatic prostate cancer received hormonal therapy (88%) in a similar proportion to an elderly cohort of American patients.^8^ Notably, of the commonest solid tumours, non-metastatic prostate cancer demonstrated the highest rate of ‘no treatment’, likely due to the use of ‘watchful-waiting’ in elderly men with prostate cancer to prevent overtreatment.^28^

For patients with non-metastatic colon cancer, Beckmann et al. demonstrated similarly high rates (83%) of surgery in an Australian cohort,^30^ when compared to our cohort (89%). Comparison of radiation therapy is difficult given that we did not differentiate between colon and rectal cancers in our study; nearly one-third of patients with rectal cancer in the aforementioned study received radiation therapy, compared to very few of those with colon cancer.^30^ Rates of systemic therapy, however, were roughly similar to data from both Australia and the US,^20, 30^ with usage increasing with increasing stage. Of all cancer types, use of targeted therapies was greatest in metastatic colorectal cancers, where EGFR (epidermal growth factor receptor) inhibitors (*e.g*. cetuximab) and anti-angiogenic agents (*e.g*. bevacizumab) are commonly used.^30^

Roughly half of ACTS participants with early-stage lung cancer received surgery, echoing US data.^20^ Notably, we did not differentiate non-small cell lung cancer and small cell lung cancer, an important delineation given that the latter is often metastatic on presentation and is therefore rarely resected. A frequency of cytotoxic chemotherapy use in patients with metastatic lung cancer receiving systemic therapy follows previously published data from Australia,^31^ the US,^20^ and the United Kingdom.^32^ Our older cohort, however, received less systemic therapy (50%) for advanced lung cancer when compared to younger cohorts; for example, Ngo et al. reported receipt of systemic therapy in 76% and 65% of patients aged <60 and 60-69 years, respectively.^31^ While only six ACTS participants (8%) received immunotherapy compared to 12% of similar patients in the US,^20^ it should be noted that immunotherapy was not routinely used during the ASPREE time period (pre-2017). Targeted therapy was as popular for advanced lung cancer in ACTS as immunotherapy; EGFR inhibitors and anti-angiogenic agents are similarly increasingly used in lung cancer.^33^

Most melanomas were treated with resection in both our cohort and worldwide.^20^ A small number of patients with non-metastatic melanoma also received immunotherapy or radiation therapy. Certainly, in the ‘real-world’, stage III melanomas may be offered adjuvant targeted therapy^34^ or immunotherapy,^35^ although these treatments were not available during the ASPREE timeframe and are less frequently used in the older population. Seventeen percent of patients with metastatic melanomas received targeted therapy; in recent years, BRAF/MEK inhibitors have shown survival benefits in BRAF-mutated disease.^36^ Similarly, immunotherapy is a common modality used in metastatic melanoma, along with radiation;^20^ roughly half of our cohort received one of the two.

We used logistic regression models to determine which factors impacted receipt of cancer treatment. In line with studies worldwide,^18-20^ participants were less likely to receive any treatment, systemic therapy, or surgery as they aged. In the case of systemic therapy, performance status, often measured using the Eastern Cooperative Oncology Group performance status (ECOG),^17^ is a key consideration in whether treatment is offered to an older patient. Similarly, pre-operative frailty and comorbidities (both of which are common in older age) are associated with post-operative morbidity and mortality and can preclude older patients from undergoing surgery.^37^ While our primary model found that female participants were more likely to receive treatment, a finding that aligns with national Australian data,^19^ this association was no longer present when sex-specific cancers were excluded. The sex difference is therefore likely driven by differing treatment patterns between the most common sex-specific cancer types; in ACTS participants with non-metastatic breast cancer, nearly all received some form of treatment, compared to only 69% of participants with non-metastatic prostate cancer. In non-sex specific cancers, we found that treatment rates were roughly similar, a finding echoed by English data.^18^ US-based participants were significantly less likely to receive most treatments compared to Australian participants. To our knowledge, there have been no head-to-head comparisons of access to cancer treatment between these two countries. Sensitivity analysis showed that this difference was partly driven by the preponderance of ethnic minorities in the US ASPREE cohort. Beyond this, however, the difference may be explained by the markedly different health systems between Australia and the US. In Australia, financial capacity is rarely a barrier to treatment, whereas high out-of-pocket expenses in the US can limit access. Notably, the small sample size of the US cancer cohort may bias the odds ratio, overestimating the effect of country of residence; therefore, the magnitude of this association should be interpreted with caution.

Several modifiable risk factors also impacted receipt of treatment. Smokers were twice as likely to not receive any treatment. Smoking status is not commonly investigated in similar studies of treatment patterns, although Ngo et al. reported no significant variation in receipt of treatment between current, former, and non-smokers in patients with lung cancer receiving systemic therapy.^31^ However, it is well-established that smoking can reduce the effectiveness of chemotherapy for lung cancer,^38^ increase symptom burden in those receiving chemotherapy and radiation therapy,^39^ and increase surgical risk,^40^ possibly impeding such patients from being offered treatment. Interestingly, current smokers were more likely to receive radiation therapy. This may be driven by smokers being less likely to receive systemic therapy or surgery due to the aforementioned risks, and instead being offered lower risk, local radiation therapy, although there is little published literature to support this hypothesis. Comorbidities may also be associated with decreased rates of treatment,^4, 18, 41^ although the evidence for whether comorbidities themselves impact receipt and efficacy of treatment is equivocal.^42, 43^ Instead, it may be that decreased performance status and/or poor baseline function secondary to comorbidities plays a greater role.

Nonetheless, ACTS participants with diabetes were less likely to receive any cancer treatment.^44^ Diabetes can increase postoperative risk,^45^ although this was not reflected in surgical treatment data in our cohort. Chronic kidney disease in our cohort was not statistically significantly associated with varying rates of any treatment. While chronic kidney disease may not impact suitability for radiation therapy or surgery, it can increase risk of toxicity and adverse reactions to systemic therapy.^46^

Notably, we did not find a statistically significant association between race, rurality, or socioeconomic status, and receipt of treatment, except for those of Asian background and receipt of radiation therapy. All of these factors have been previously found to be associated with varying rates of cancer treatment,^4, 19, 47^ although in Australia, remoteness and socioeconomic status do not appear to impact receipt of treatment.^19^ Note that our data do not provide information on whether these factors affect ease of access to cancer treatment or timeliness of treatment. Nonetheless, our findings may be explained by variation between cohorts, or that our sample sizes for each stratum were relatively small. Notably, while our regression models did not reveal significant associations between these factors, the descriptive data did highlight some differences. For example, 83% of Caucasian participants received treatment compared with 75% of African American participants. Similarly, we saw that treatment rates decreased with increasing rurality, although the majority of those living in inner and outer regional Australia still received treatment (83% and 79%, respectively).

Furthermore, the impact of these factors is likely to vary between countries depending on health systems, level of reimbursement, and location-based access to treatment. Alternatively, it has been suggested that these differences may be explained by different cancer types or characteristics seen in different races or socioeconomic groups.^18^ We also found no significant impact of frailty on receipt of cancer treatment, although due to ASPREE’s strict inclusion criteria, few participants were frail at enrolment. Furthermore, frailty was only measured at baseline in ASPREE; it is likely that over the course of the study more participants developed frailty. Similarly, we found no statistically significant impact of BMI on receipt of treatment, despite previous literature describing that high (but not excessively high) BMI can improve outcomes for both systemic therapy^31^ and surgery.^48^ For both frailty and BMI, however, the point estimates for odds ratios were less and greater than 1.00 respectively and may therefore partly reflect a need for a larger sample.

This analysis has several strengths. Firstly, the large cohort of the ASPREE trial allowed for a relatively large number of cancer events to be captured with a reasonable spread of cancer types and stages, along with a reasonably representative sample. However, the limited sample size in the US, and modest number of participants within each cancer type, limits our ability to investigate heterogeneity in treatment patterns across cancer types and countries. The adjudication of cancer cases allowed for accurate classification of cancer events. The in-depth baseline data captured by ASPREE was also a significant strength and allowed us to examine the relationship of various participant characteristics and receipt of treatment. Finally, cancer treatment data were available for a significant proportion (98%) of the ACTS cohort, allowing for a relatively robust dataset. The rigorous inclusion criteria for ASPREE requiring participants to be ‘healthy’ however, may be a limitation in our analysis. These participants are typically more likely to receive treatment due to greater baseline functional status, thereby overestimating treatment rates. A lack of specific treatment data also prevents more detailed analysis of treatment patterns. We also did not collect data on treatment intent; as a result, we were unable to differentiate between rates of curative versus palliative treatment.

Despite cancer treatment typically having a positive impact on cancer-related mortality and recurrence, both cancer and cancer treatment are thought to have the potential to accelerate adverse ageing outcomes in cancer survivors.^49, 50^ Therefore, the ACTS substudy data will be used in subsequent analyses to explore long-term outcomes of older patients with cancer including the impact of cancer and cancer treatment on cognitive decline and dementia, cardiovascular disease, and functional decline.

## Conclusion

This study is one of the first to describe cancer treatment patterns and factors associated with receipt of treatment across several cancer types in older adults in Australia and the US. In our cohort, most older adults with cancer received some form of cancer treatment, typically only one type, and most commonly being surgery. Younger and female participants more frequently received cancer treatment than older participants, although the sex association was predominantly driven by differing treatment rates for sex-specific cancers. Residence in the US, current smoking status, and diabetes, reduced the likelihood of receiving any treatment.

## Data Availability

Data are available from the corresponding author upon reasonable request and approval of the ASPREE Steering Committee.

## Abbreviations

(ASPREE): ASPirin in Reducing Events in the Elderly
(ACTS): ASPREE Cancer Treatment Substudy
(BMI): Body Mass Index
CI: (Confidence Interval)
(ECOG): Eastern Cooperative Oncology Group
(EGFR): Epidermal growth factor receptor
(IRSAD): Index of Relative Socio-economic Advantage and Disadvantage
(OR): Odds Ratio
(SEER): Surveillance, Epidemiology, and End Results
(US): United States

## Ethics statement

Ethics approval for the ASPREE trial was received from each participating centre in Australia and the United States. Additionally, the National Institute on Aging, the principal funder of the trial, along with an independent board assigned to monitor data and safety, reviewed the data collected by the trial biannually. Ethics approval for this substudy (Project 390/20) was granted by the Alfred Hospital Ethics Committee.

## Author contributions

JM: conceptualisation, data curation, formal analysis, investigation, methodology, project administration, visualisation, writing – original draft. EW: conceptualisation, funding acquisition, investigation, methodology, project administration, supervision, writing – original draft. JZ: conceptualisation, funding acquisition, investigation, methodology, supervision, validation, writing – original draft. AH: conceptualisation, investigation, methodology, supervision, validation. GP: data curation, formal analysis, methodology, visualisation. GvL: conceptualisation, investigation, methodology, supervision, validation, writing – original draft. PG: investigation, validation. WB: investigation, validation. JT: investigation, validation. VM: investigation, validation. JLM: investigation, validation. JMcN: funding acquisition, investigation. RLW: funding acquisition, data acquisition, project management, investigation. SO: conceptualisation, data curation, formal analysis, investigation, methodology, project administration, supervision, writing – original draft.

All authors were involved in writing (review and editing). The work reported in the paper has been performed by the authors, unless clearly specified in the text.

## Footnotes

a This group includes treatments including regional chemotherapy (e.g. transarterial chemoembolisation) and regional immunotherapy (e.g. intravesical BCG). Supplementary Table 1 contains the full definition for regional therapy.

b Note that the models for each modality are independent of one another and therefore ORs cannot be compared between modalities.

## References

1. Sung H, Ferlay J, Siegel RL, Laversanne M, Soerjomataram I, Jemal A, Bray F. Global Cancer Statistics 2020: GLOBOCAN Estimates of Incidence and Mortality Worldwide for 36 Cancers in 185 Countries. CA Cancer J Clin. 2021;71(3):209–49. https://doi.org/10.3322/caac.21660.

2. Bluethmann SM, Mariotto AB, Rowland JH. Anticipating the “Silver Tsunami”: Prevalence Trajectories and Comorbidity Burden among Older Cancer Survivors in the United States. Cancer Epidemiol Biomarkers Prev. 2016;25(7):1029–36. https://doi.org/10.1158/1055-9965.EPI-16-0133.

3. Armenian SH, Gibson CJ, Rockne RC, Ness KK. Premature Aging in Young Cancer Survivors. J Natl Cancer Inst. 2019;111(3):226–32. https://doi.org/10.1093/jnci/djy229.

4. Morimoto L, Coalson J, Mowat F, O’Malley C. Factors affecting receipt of chemotherapy in women with breast cancer. Int J Womens Health. 2010;2:107–22. https://doi.org/10.2147/ijwh.s9125.

5. Shavers VL, Brown ML. Racial and ethnic disparities in the receipt of cancer treatment. J Natl Cancer Inst. 2002;94(5):334–57. https://doi.org/10.1093/jnci/94.5.334.

6. Given B, Given CW. Older adults and cancer treatment. Cancer. 2008;113(12 Suppl):3505–11. https://doi.org/10.1002/cncr.23939.

7. Sedrak MS, Freedman RA, Cohen HJ, Muss HB, Jatoi A, Klepin HD, Wildes TM, Le-Rademacher JG, Kimmick GG, Tew WP, et al. Older adult participation in cancer clinical trials: A systematic review of barriers and interventions. CA Cancer J Clin. 2021;71(1):78–92. https://doi.org/10.3322/caac.21638.

8. Bunniran S, Xu Y, Molife C, Nair R, Zhu YE, Carter GC, Sheth S, Hess LM. Treatment patterns for patients with advanced/metastatic cancers by site of care. Am J Manag Care. 2021;27(4):e105–e13. https://doi.org/10.37765/ajmc.2021.88619.

9. Kurosky SK, Mitra D, Zanotti G, Kaye JA. Treatment Patterns and Outcomes of Patients With Metastatic ER(+)/HER-2(-) Breast Cancer: A Multicountry Retrospective Medical Record Review. Clin Breast Cancer. 2018;18(4):e529–e38. https://doi.org/10.1016/j.clbc.2017.10.008.

10. McNeil JJ, Woods RL, Nelson MR, Murray AM, Reid CM, Kirpach B, Storey E, Shah RC, Wolfe RS, Tonkin AM, et al. Baseline Characteristics of Participants in the ASPREE (ASPirin in Reducing Events in the Elderly) Study. J Gerontol A Biol Sci Med Sci. 2017;72(11):1586–93. https://doi.org/10.1093/gerona/glw342.

11. Group AI. Study design of ASPirin in Reducing Events in the Elderly (ASPREE): a randomized, controlled trial. Contemp Clin Trials. 2013;36(2):555–64. https://doi.org/10.1016/j.cct.2013.09.014.

12. McNeil JJ, Nelson MR, Woods RL, Lockery JE, Wolfe R, Reid CM, Kirpach B, Shah RC, Ives DG, Storey E, et al. Effect of Aspirin on All-Cause Mortality in the Healthy Elderly. N Engl J Med. 2018;379(16):1519–28. https://doi.org/10.1056/NEJMoa1803955.

13. McNeil JJ, Wolfe R, Woods RL, Tonkin AM, Donnan GA, Nelson MR, Reid CM, Lockery JE, Kirpach B, Storey E, et al. Effect of Aspirin on Cardiovascular Events and Bleeding in the Healthy Elderly. N Engl J Med. 2018;379(16):1509–18. https://doi.org/10.1056/NEJMoa1805819.

14. McNeil JJ, Woods RL, Nelson MR, Reid CM, Kirpach B, Wolfe R, Storey E, Shah RC, Lockery JE, Tonkin AM, et al. Effect of Aspirin on Disability-free Survival in the Healthy Elderly. N Engl J Med. 2018;379(16):1499–508. https://doi.org/10.1056/NEJMoa1800722.

15. Orchard SG, Lockery JE, Gibbs P, Polekhina G, Wolfe R, Zalcberg J, Haydon A, McNeil JJ, Nelson MR, Reid CM, et al. Cancer history and risk factors in healthy older people enrolling in the ASPREE clinical trial. Contemp Clin Trials. 2020;96:106095. https://doi.org/10.1016/j.cct.2020.106095.

16. McNeil JJ, Gibbs P, Orchard SG, Lockery JE, Bernstein WB, Cao Y, Ford L, Haydon A, Kirpach B, Macrae F, et al. Effect of Aspirin on Cancer Incidence and Mortality in Older Adults. J Natl Cancer Inst. 2021;113(3):258–65. https://doi.org/10.1093/jnci/djaa114.

17. 1. Schnipper LE, Smith TJ, Raghavan D, Blayney DW, Ganz PA, Mulvey TM, Wollins DS. American Society of Clinical Oncology identifies five key opportunities to improve care and reduce costs: the top five list for oncology. J Clin Oncol. 2012;30(14):1715–24. https://doi.org/10.1200/jco.2012.42.8375.

18. Chemotherapy, Radiotherapy and Surgical Tumour Resections in England. Public Health England; 2020.

19. Cancer control continuum | Treatment: Cancer Australia; 2017 [cited 23 May]. Available from: https://ncci.canceraustralia.gov.au/treatment.

20. Miller KD, Nogueira L, Mariotto AB, Rowland JH, Yabroff KR, Alfano CM, Jemal A, Kramer JL, Siegel RL. Cancer treatment and survivorship statistics, 2019. CA Cancer J Clin. 2019;69(5):363–85. https://doi.org/10.3322/caac.21565.

21. Lang K, Earle CC, Foster T, Dixon D, Van Gool R, Menzin J. Trends in the treatment of acute myeloid leukaemia in the elderly. Drugs Aging. 2005;22(11):943–55. https://doi.org/10.2165/00002512-200522110-00004.

22. Medeiros BC, Satram-Hoang S, Hurst D, Hoang KQ, Momin F, Reyes C. Big data analysis of treatment patterns and outcomes among elderly acute myeloid leukemia patients in the United States. Ann Hematol. 2015;94(7):1127–38. https://doi.org/10.1007/s00277-015-2351-x.

23. Hamlin PA, Satram-Hoang S, Reyes C, Hoang KQ, Guduru SR, Skettino S. Treatment patterns and comparative effectiveness in elderly diffuse large B-cell lymphoma patients: a surveillance, epidemiology, and end results-medicare analysis. Oncologist. 2014;19(12):1249–57. https://doi.org/10.1634/theoncologist.2014-0113.

24. Hallek M, Cheson BD, Catovsky D, Caligaris-Cappio F, Dighiero G, Dohner H, Hillmen P, Keating M, Montserrat E, Chiorazzi N, et al. iwCLL guidelines for diagnosis, indications for treatment, response assessment, and supportive management of CLL. Blood. 2018;131(25):2745–60. https://doi.org/10.1182/blood-2017-09-806398.

25. Merie R, Shafiq J, Soon PS, Delaney GP. Surgical and radiotherapy patterns of care in the management of breast cancer in NSW and ACT Australia. J Med Imaging Radiat Oncol. 2022;66(3):442–54. https://doi.org/10.1111/1754-9485.13357.

26. Hurria A, Leung D, Trainor K, Borgen P, Norton L, Hudis C. Factors influencing treatment patterns of breast cancer patients age 75 and older. Crit Rev Oncol Hematol. 2003;46(2):121–6. https://doi.org/10.1016/s1040-8428(02)00133-6.

27. Burt LM, Shrieve DC, Tward JD. Factors influencing prostate cancer patterns of care: An analysis of treatment variation using the SEER database. Adv Radiat Oncol. 2018;3(2):170–80. https://doi.org/10.1016/j.adro.2017.12.008.

28. Wang LL, Begashaw K, Evans M, Earnest A, Evans SM, Millar JL, Murphy DG, Moon D. Patterns of care and outcomes for men diagnosed with prostate cancer in Victoria: an update. ANZ J Surg. 2018;88(10):1037–42. https://doi.org/10.1111/ans.14722.

29. Hamdy FC, Donovan JL, Lane JA, Mason M, Metcalfe C, Holding P, Davis M, Peters TJ, Turner EL, Martin RM, et al. 10-Year Outcomes after Monitoring, Surgery, or Radiotherapy for Localized Prostate Cancer. N Engl J Med. 2016;375(15):1415–24. https://doi.org/10.1056/NEJMoa1606220.

30. Beckmann KR, Bennett A, Young GP, Roder DM. Treatment patterns among colorectal cancer patients in South Australia: a demonstration of the utility of population-based data linkage. J Eval Clin Pract. 2014;20(4):467–77. https://doi.org/10.1111/jep.12183.

31. Ngo P, Goldsbury DE, Karikios D, Yap S, Yap ML, Egger S, O’Connell DL, Ball D, Fong KM, Pavlakis N, et al. Lung cancer treatment patterns and factors relating to systemic therapy use in Australia. Asia-Pacific Journal of Clinical Oncology.n/a(n/a). https://doi.org/https://doi.org/10.1111/ajco.13637.

32. Lester J, Escriu C, Khan S, Hudson E, Mansy T, Conn A, Chan S, Powell C, Brock J, Conibear J, et al. Retrospective analysis of real-world treatment patterns and clinical outcomes in patients with advanced non-small cell lung cancer starting first-line systemic therapy in the United Kingdom. BMC Cancer. 2021;21(1):515. https://doi.org/10.1186/s12885-021-08096-w.

33. Majeed U, Manochakian R, Zhao Y, Lou Y. Targeted therapy in advanced non-small cell lung cancer: current advances and future trends. J Hematol Oncol. 2021;14(1):108. https://doi.org/10.1186/s13045-021-01121-2.

34. Long GV, Hauschild A, Santinami M, Atkinson V, Mandala M, Chiarion-Sileni V, Larkin J, Nyakas M, Dutriaux C, Haydon A, et al. Adjuvant Dabrafenib plus Trametinib in Stage III BRAF-Mutated Melanoma. N Engl J Med. 2017;377(19):1813–23. https://doi.org/10.1056/NEJMoa1708539.

35. Eggermont AMM, Blank CU, Mandala M, Long GV, Atkinson V, Dalle S, Haydon A, Lichinitser M, Khattak A, Carlino MS, et al. Adjuvant Pembrolizumab versus Placebo in Resected Stage III Melanoma. N Engl J Med. 2018;378(19):1789–801. https://doi.org/10.1056/NEJMoa1802357.

36. Robert C, Karaszewska B, Schachter J, Rutkowski P, Mackiewicz A, Stroiakovski D, Lichinitser M, Dummer R, Grange F, Mortier L, et al. Improved overall survival in melanoma with combined dabrafenib and trametinib. N Engl J Med. 2015;372(1):30–9. https://doi.org/10.1056/NEJMoa1412690.

37. Partridge JS, Harari D, Dhesi JK. Frailty in the older surgical patient: a review. Age Ageing. 2012;41(2):142–7. https://doi.org/10.1093/ageing/afr182.

38. Condoluci A, Mazzara C, Zoccoli A, Pezzuto A, Tonini G. Impact of smoking on lung cancer treatment effectiveness: a review. Future Oncol. 2016;12(18):2149–61. https://doi.org/10.2217/fon-2015-0055.

39. Peppone LJ, Mustian KM, Morrow GR, Dozier AM, Ossip DJ, Janelsins MC, Sprod LK, McIntosh S. The effect of cigarette smoking on cancer treatment-related side effects. Oncologist. 2011;16(12):1784–92. https://doi.org/10.1634/theoncologist.2011-0169.

40. Hawn MT, Houston TK, Campagna EJ, Graham LA, Singh J, Bishop M, Henderson WG. The attributable risk of smoking on surgical complications. Ann Surg. 2011;254(6):914–20. https://doi.org/10.1097/SLA.0b013e31822d7f81.

41. Hall WH, Jani AB, Ryu JK, Narayan S, Vijayakumar S. The impact of age and comorbidity on survival outcomes and treatment patterns in prostate cancer. Prostate Cancer Prostatic Dis. 2005;8(1):22–30. https://doi.org/10.1038/sj.pcan.4500772.

42. Mellemgaard A, Luchtenborg M, Iachina M, Jakobsen E, Green A, Krasnik M, Moller H. Role of comorbidity on survival after radiotherapy and chemotherapy for nonsurgically treated lung cancer. J Thorac Oncol. 2015;10(2):272–9. https://doi.org/10.1097/JTO.0000000000000416.

43. Lee L, Cheung WY, Atkinson E, Krzyzanowska MK. Impact of comorbidity on chemotherapy use and outcomes in solid tumors: a systematic review. J Clin Oncol. 2011;29(1):106–17. https://doi.org/10.1200/JCO.2010.31.3049.

44. Gerards MC, van der Velden DL, Baars JW, Brandjes DPM, Hoekstra JBL, Vriesendorp TM, Gerdes VEA. Impact of hyperglycemia on the efficacy of chemotherapy-A systematic review of preclinical studies. Crit Rev Oncol Hematol. 2017;113:235–41. https://doi.org/10.1016/j.critrevonc.2017.03.007.

45. Wang J, Chen K, Li X, Jin X, An P, Fang Y, Mu Y. Postoperative adverse events in patients with diabetes undergoing orthopedic and general surgery. Medicine (Baltimore). 2019;98(14):e15089. https://doi.org/10.1097/MD.0000000000015089.

46. Bednarek A, Mykala-Ciesla J, Pogoda K, Jagiello-Gruszfeld A, Kunkiel M, Winder M, Chudek J. Limitations of Systemic Oncological Therapy in Breast Cancer Patients with Chronic Kidney Disease. J Oncol. 2020;2020:7267083. https://doi.org/10.1155/2020/7267083.

47. Hendren S, Chin N, Fisher S, Winters P, Griggs J, Mohile S, Fiscella K. Patients’ barriers to receipt of cancer care, and factors associated with needing more assistance from a patient navigator. J Natl Med Assoc. 2011;103(8):701–10. https://doi.org/10.1016/s0027-9684(15)30409-0.

48. Ri M, Aikou S, Seto Y. Obesity as a surgical risk factor. Ann Gastroenterol Surg. 2018;2(1):13–21. https://doi.org/10.1002/ags3.12049.

49. Guida JL, Ahles TA, Belsky D, Campisi J, Cohen HJ, DeGregori J, Fuldner R, Ferrucci L, Gallicchio L, Gavrilov L, et al. Measuring Aging and Identifying Aging Phenotypes in Cancer Survivors. J Natl Cancer Inst. 2019;111(12):1245–54. https://doi.org/10.1093/jnci/djz136.

50. Muhandiramge J, Orchard S, Haydon A, Zalcberg J. The acceleration of ageing in older patients with cancer. J Geriatr Oncol. 2020. https://doi.org/10.1016/j.jgo.2020.09.010.

