## Supplementary for "Cancer treatment patterns and factors affecting receipt of treatment in older adults: results from the ASPREE Cancer Treatment Substudy (ACTS)"

*Supplementary Figure 1. Flow diagram of inclusion for the ASPREE Cancer Treatment Substudy (ACTS).*

**ASPREE cohort**

n = 19,114

**Post-randomisation cancer diagnosis**

n = 1,933

**Excluded participants without a post-randomisation cancer diagnosis**

n = 17,181

**Cancer with complete treatment data (ACTS cohort)**

n = 1,893

**Excluded participants with insufficient treatment data**

n = 40

**Cancer with cancer treatment**

n = 1,569

**Cancer with no cancer treatment**

n = 324

*Supplementary Table 1. Coding criteria for cancer treatment data, stratified by treatment modality.*

| **Type of cancer treatment data** | **Criteria** |
| --- | --- |
| Systemic therapy   - Cytotoxic chemotherapy - Hormonal therapy - Targeted therapy - Immunotherapy | *Inclusion*   - Any systemic therapy including, but not limited to, cytotoxic chemotherapy, hormonal therapy, targeted therapy, and immunotherapy   *Exclusion*   - Regional chemotherapy *e.g.* transarterial chemoembolisation, hypothermic intraperitoneal chemotherapy, intravesical mitomycin c - Regional immunotherapy *e.g.* intravesical BCG (Bacillus Calmette-Guerin) immunotherapy (early-stage bladder cancer)   *Miscellaneous*   - Arsenic trioxide + all-trans retinoic acid for acute promyelocytic leukaemia should be coded as targeted therapy |
| Radiation therapy | *Inclusion*   - Any radiation therapy procedure used in the treatment of a primary tumour or its metastases   *Exclusions*   - None   *Miscellaneous*   - Radioactive iodine for thyroid cancer should be coded as radiation therapy |
| Surgery | *Inclusion*   - Any operation that has been undertaken to remove a tumour *e.g.* anterior resection for colorectal cancer, mastectomy for breast cancer, wide local excisions of melanoma, endoscopic resection of a known tumour - Any operation that has been undertaken to debulk the tumour - Any operation related to the treatment of the tumour or its associated symptoms *e.g.* video-assisted thoracoscopic pleurodesis for pleural effusion secondary to lung cancer, transurethral resection of the prostate for prostatomegaly secondary to prostate cancer, bypass surgery for pancreatic surgery   *Exclusion*   - Endoscopic procedures without intent for curative resection of a tumour *e.g.* colonoscopy, cystoscopy, bronchoscopy - Biopsies and other diagnostic procedures *e.g*. excision biopsies for melanoma or breast cancer - Procedures in which a cancer was incidentally discovered *e.g*. diagnosis of prostate cancer on transurethral resection of the prostate for benign prostatic hyperplasia |
| Regional therapy | *Inclusion*   - Regional chemotherapy *e.g.* transarterial chemoembolisation, hypothermic intraperitoneal chemotherapy, intravesical mitomycin c - Regional immunotherapy for confirmed cancers *e.g*. intravesical BCG (Bacillus Calmette-Guerin) immunotherapy   *Exclusion*   - Regional therapy for in-situ cancers *e.g.* intravesical BCG for bladder carcinomas in-situ |

*Supplementary Table 2. Cancer treatment modalities received for other incident post-randomisation cancers during ASPREE, stratified by cancer type and metastatic status, and cause of death and time from diagnosis to death.*

|  |  | **Systemic Therapy** | | | | |  |  |
| --- | --- | --- | --- | --- | --- | --- | --- | --- |
|  | **Any treatment**  **(n=1,569)** | **Any systemic therapy (n=869)** | **Chemotherapy (n=537)** | **Hormonal therapy (n=351)** | **Targeted therapy (n=85)** | **Immunotherapy (n=31)** | **Radiation therapy (n=544)** | **Surgery (n=1,029)** |
| **Non-metastatic solid tumours** | | | | | | | | |
| Bladder (n=59) | 48 (81%) | 15 (25%) | 15 (25%) | - | - | 1 (2%) | 9 (15%) | 32 (54%) |
| Brain (n=25) | 17 (68%) | 8 (32%) | 8 (32%) | - | - | - | 10 (40%) | 16 (64%) |
| Cervical (n=3) | 3 (100%) | 3 (100%) | 3 (100%) | - | - | - | 3 (100%) | - |
| Gallbladder or bile duct (n=11) | 6 (55%) | 4 (36%) | 4 (36%) | - | - | - | 2 (18%) | 3 (27%) |
| Kidney (n=25) | 24 (96%) | 1 (4%) | - | - | 1 (4%) | - | 1 (4%) | 24 (96%) |
| Liver (n=3) | 2 (67%) | 2 (67%) | - | - | 2 (67%) | 1 (33%) | 1 (33%) | 1 (33%) |
| Mesothelioma (n=14) | 12 (86%) | 6 (43%) | 6 (43%) | - | - | - | 3 (21%) | 8 (57%) |
| Oesophageal (n=14) | 13 (93%) | 8 (57%) | 8 (57%) | - | - | - | 11 (79%) | 5 (36%) |
| Ovary or endometrium (n=39) | 36 (92%) | 12 (31%) | 11 (28%) | 1 (3%) | - | - | 15 (38%) | 35 (90%) |
| Pancreas (n=31) | 23 (74%) | 14 (45%) | 14 (45%) | - | - | - | 2 (6%) | 18 (58%) |
| Stomach (n=21) | 17 (81%) | 8 (38%) | 8 (38%) | - | - | - | 5 (24%) | 16 (76%) |
| Thyroid (n=10) | 10 (100%) | - | - | - | - | - | 6 (60%) | 9 (90%) |
| **Metastatic solid tumours** | | | | | | | | |
| Bladder (n=10) | 7 (70%) | 4 (40%) | 4 (40%) | - | - | - | 3 (30%) | 2 (20%) |
| Brain (n=0) | N/A | N/A | N/A | N/A | N/A | N/A | N/A | N/A |
| Cervical (n=0) | N/A | N/A | N/A | N/A | N/A | N/A | N/A | N/A |
| Gallbladder or bile duct (n=10) | 5 (50%) | 5 (50%) | 5 (50%) | - | - | - | - | 1 (10%) |
| Kidney (n=10) | 7 (70%) | 3 (30%) | 2 (20%) | - | 1 (10%) | - | 4 (40%) | 4 (40%) |
| Liver (n=4) | 3 (75%) | 1 (25%) | - | - | 1 (25%) | - | 1 (25%) | 1 (25%) |
| Mesothelioma (n=1) | 1 (100%) | - | - | - | - | - | - | 1 (100%) |
| Oesophageal (n=13) | 11 (85%) | 9 (69%) | 9 (69%) | - | - | - | 9 (69%) | - |
| Ovary or endometrium (n=34) | 28 (82%) | 24 (71%) | 23 (68%) | 2 (6%) | 3 (9%) | - | 2 (6%) | 18 (53%) |
| Pancreas (n=33) | 20 (61%) | 16 (48%) | 16 (48%) | - | - | - | 2 (6%) | 7 (21%) |
| Stomach (n=8) | 6 (75%) | 2 (25%) | 2 (25%) | - | - | - | 3 (38%) | 1 (13%) |
| Thyroid (n=0) | N/A | N/A | N/A | N/A | N/A | N/A | N/A | N/A |
| Percentages represent the proportion of participants who received cancer treatment relative to the first column. For example, 81% of participants with bladder cancers received any cancer treatment.  “-“ indicates that no one (0 participants) in this group received a particular treatment modality. | | | | | | | | |

*Supplementary Table 3. Cancer treatment modalities received for common non-sex specific incident post-randomisation cancers during ASPREE, stratified by cancer type and sex (F = female, M = male).*

|  |  | | **Systemic Therapy** | | | | | | | |  | |  | |
| --- | --- | --- | --- | --- | --- | --- | --- | --- | --- | --- | --- | --- | --- | --- |
|  | **Any treatment**  **(n=1,569)** | | **Any systemic therapy (n=869)** | | **Chemotherapy (n=537)** | | **Targeted therapy (n=85)** | | **Immunotherapy (n=31)** | | **Radiation therapy (n=544)** | | **Surgery (n=1,029)** | |
|  | *F* | *M* | *F* | *M* | *F* | *M* | *F* | *M* | *F* | *M* | *F* | *M* | *F* | *M* |
| **Non-metastatic solid tumours** | | | | | | | | | | | | | | |
| Colorectal (n_F_=98; n_m_=108) | 90 (92%) | 101 (94%) | 30 (31%) | 41 (38%) | 30 (31%) | 41 (38%) | 1 (1%) | 3 (3%) | - | - | 7 (7%) | 13 (12%) | 98 (100%) | 98 (91%) |
| Lung (n_F_=31; n_m_=44) | 29 (94%) | 36 (82%) | 10 (32%) | 10 (23%) | 9 (29%) | 9 (20%) | - | 1 (2%) | 2 (6%) | - | 14 (45%) | 19 (43%) | 16 (52%) | 22 (50%) |
| Melanoma (n_F_=60; n_m_=100) | 57 (95%) | 94 (94%) | 3 (5%) | 2 (2%) | - | - | - | - | 3 (5%) | 2 (2%) | 1 (2%) | 4 (4%) | 55 (92%) | 91 (91%) |
| **Metastatic solid tumours** | | | | | | | | | | | | | | |
| Colorectal (n_F_=26; n_m_=30) | 23 (88%) | 27 (90%) | 20 (77%) | 22 (73%) | 20 (77%) | 22 (73%) | 12 (46%) | 8 (27%) | - | - | 4 (15%) | 7 (23%) | 14 (54%) | 21 (70%) |
| Lung (n_F_=26; n_m_=51) | 20 (77%) | 39 (76%) | 12 (46%) | 27 (53%) | 9 (35%) | 27 (53%) | 3 (12%) | 3 (6%) | 2 (8%) | 4 (8%) | 16 (62%) | 24 (47%) | 5 (19%) | 10 (20%) |
| Melanoma (n_F_=10; n_m_=18) | 8 (80%) | 15 (83%) | 3 (30%) | 12 (67%) | 1 (10%) | 1 (6%) | 1 (10%) | 4 (22%) | 1 (10%) | 10 (56%) | 3 (30%) | 7 (39%) | 5 (50%) | 11 (61%) |
| **Haematological malignancy** (n_F_=97; n_m_=88) | 64 (66%) | 48 (55%) | 58 (60%) | 44 (50%) | 54 (56%) | 43 (49%) | 10 (10%) | 9 (10%) | - | 2 (2%) | 10 (10%) | 6 (7%) | 6 (6%) | 4 (5%) |
| Percentages represent the proportion of participants who received cancer treatment relative to the first column. For example, 92% of female participants with non-metastatic colorectal cancers received any cancer treatment.  “-“ indicates that no one (0 participants) in this group received a particular treatment modality. | | | | | | | | | | | | | | |

*Supplementary Table 4. Cancer treatment modalities received for common incident post-randomisation cancers during ASPREE, stratified by cancer type and country of residence (Aus = Australia, US = United States).*

|  |  | | **Systemic Therapy** | | | | | | | | | |  | |  | |
| --- | --- | --- | --- | --- | --- | --- | --- | --- | --- | --- | --- | --- | --- | --- | --- | --- |
|  | **Any treatment**  **(n=1,569)** | | **Any systemic therapy (n=869)** | | **Chemotherapy (n=537)** | | **Hormonal therapy (n=351)** | | **Targeted therapy (n=85)** | | **Immunotherapy (n=31)** | | **Radiation therapy (n=544)** | | **Surgery (n=1,029)** | |
|  | *Aus* | *US* | *Aus* | *US* | *Aus* | *US* | *Aus* | *US* | *Aus* | *US* | *Aus* | *US* | *Aus* | *US* | *Aus* | *US* |
| **All cancers (**n_Aus_=1,758; n_US_=175) | 1,444 (82%) | 125 (71%) | 799 (45%) | 70 (40%) | 501 (28%) | 36 (21%) | 317 (18%) | 34 (19%) | 82 (5%) | 3 (2%) | 27 (2%) | 4 (2%) | 512 (29%) | 32 (18%) | 944 (54%) | 85 (49%) |
| **Non-metastatic solid tumours** | | | | | | | | | | | | | | | | |
| Breast (n_Aus_=181; n_US_=30) | 181 (100%) | 29 (97%) | 150 (83%) | 21 (70%) | 46 (25%) | 3 (10%) | 129 (71%) | 20 (67%) | 13 (7%) | - | - | - | 97 (54%) | 7 (23%) | 179 (99%) | 27 (90%) |
| Colon/rectum (n_Aus_=198; n_US_=8) | 185 (93%) | 6 (75%) | 68 (34%) | 3 (38%) | 68 (34%) | 3 (38%) | N/A | N/A | 4 (2%) | - | - | - | 19 (10%) | 1 (13%) | 182 (92%) | 5 (63%) |
| Lung (n_Aus_=68; n_US_=7) | 60 (88%) | 5 (71%) | 20 (29%) | - | 18 (26%) | - | N/A | N/A | 1 (1%) | - | 2 (3%) | - | 33 (49%) | - | 33 (49%) | 5 (71%) |
| Melanoma (n_Aus_=154; n_US_=6) | 149 (97%) | 2 (33%) | 4 (3%) | 1 (17%) | - | - | N/A | N/A | - | - | 4 (3%) | 1 (17%) | 5 (3%) | - | 145 (94%) | 1 (17%) |
| Prostate (n_Aus_=248; n_US_=18) | 182 (73%) | 9 (50%) | 94 (38%) | 4 (22%) | 5 (2%) | - | 92 (37%) | 4 (22%) | - | - | - | - | 102 (41%) | 4 (22%) | 79 (32%) | 4 (22%) |
| **Metastatic solid tumours** | | | | | | | | | | | | | | | | |
| Breast (n_Aus_=27; n_US_=5) | 24 (89%) | 4 (80%) | 22 (81%) | 4 (80%) | 8 (30%) | 2 (40%) | 17 (63%) | 3 (60%) | 4 (15%) | - | - | - | 12 (44%) | 3 (60%) | 7 (26%) | 1 (20%) |
| Colon/rectum (n_Aus_=54; n_US_=2) | 49 (91%) | 1 (50%) | 41 (76%) | 1 (50%) | 41 (76%) | 1 (50%) | N/A | N/A | 20 (37%) | - | - | - | 11 (20%) | - | 35 (65%) | - |
| Lung (n_Aus_=68; n_US_=9) | 54 (79%) | 5 (56%) | 36 (53%) | 3 (33%) | 33 (49%) | 3 (33%) | N/A | N/A | 6 (9%) | - | 4 (6%) | 2 (22%) | 36 (53%) | 4 (44%) | 15 (22%) | - |
| Melanoma (n_Aus_=25; n_US_=3) | 21 (84%) | 2 (67%) | 14 (56%) | 1 (33%) | 1 (4%) | 1 (33%) | N/A | N/A | 5 (20%) | - | 11 (44%) | - | 9 (36%) | 1 (33%) | 15 (60%) | 1 (33%) |
| Prostate (n_Aus_=87; n_US_=11) | 80 (92%) | 8 (73%) | 78 (90%) | 8 (73%) | 24 (28%) | 1 (9%) | 76 (87%) | 7 (64%) | - | - | - | - | 31 (36%) | 4 (36%) | 16 (18%) | 2 (18%) |
| **Haematological malignancy** (n_Aus_=164; n_US_=21) | 103 (63%) | 9 (43%) | 94 (57%) | 8 (38%) | 90 (55%) | 7 (33%) | N/A | N/A | 18 (11%) | 1 (5%) | 2 (1%) | - | 14 (9%) | 2 (10%) | 10 (6%) | - |
| NB: Percentages represent the proportion of participants who received cancer treatment relative to the first column. For example, 100% of Australian participants with non-metastatic breast cancers received any cancer treatment.  “-“ indicates that no one (0 participants) in this group received a particular treatment modality. | | | | | | | | | | | | | | | | |
